# Vegetable intake and metabolic risk factors: A Mendelian randomization study

**DOI:** 10.1101/2022.03.21.22272720

**Authors:** Q Feng, et al.

## Abstract

**Background:** The associations between vegetable intake and metabolic risk factors remain inconsistent. This study was aimed to investigate the association between cooked and raw vegetable intake with serum lipids, body mass index (BMI), blood pressure and glycemic traits.

**Methods:** This was a two-sample Mendelian randomization study. Nine and 19 genetic variants were identified from genome-wide association studies (GWAS) as instrumental variables for cooked and raw vegetable intake. Summary-level statistics were used from GWAS of total cholesterol, triglycerides, high-density lipoprotein, low-density lipoprotein, systolic blood pressure, diastolic blood pressure, BMI, fasting glucose, fasting insulin, glycated haemoglobin and 2-hour glucose after oral glucose tolerance test. Multivariable MR with inverse-variance weighted method was performed as primary analysis, while median-based method and MR-Egger method were performed as sensitivity analyses.

**Results:** Vegetable intake was not associated with total cholesterol (−0.06 [−0.30, 0.18] and −0.02 [−0.20, 0.16] for each serving increase of cooked and raw vegetable intake) and triglyceride (−0.12 [−0.36, 0.13] and 0.03 [−0.15, 0.22]). Null evidence was observed for associations between vegetable intake and lipoproteins, blood pressure, BMI, insulin and glycemic measures. Sensitivity analyses generated similar null associations.

**Conclusions:** We found null association between vegetable intake and metabolic risk factors. This is consistent with previous MR findings of null associations between diet-derived antioxidants and metabolic risk factors.

## 1. Introduction

Metabolic risk factors, including obesity, hypertension, abnormal glycemic traits and dyslipidemia, are the most prevalent risk factors for non-communicable diseases [1]. High levels of systolic blood pressure (SBP), fasting glucose (FG), body mass index (BMI) and low-density lipoprotein cholesterol (LDL) consist of 9.3%, 6.8%, 6.3% and 3.9% of global disability-adjusted life-years [1], respectively. They have been ranked among the top 10 risk factors for cardiovascular disease burden in adults [2].

Sufficient vegetable consumption has been widely recommended to reduce risks of non-communicable diseases [3]. The associations between vegetable intake and metabolic risk factors have been investigated by previous studies, however the results remain unclear [4]. An early meta-analysis of randomized controlled trials reported reduced body weight following increased vegetable and fruit intake [5], which was conflicting with more recent evidence showing null effects on body weight [6–8]. A systematic review of observational studies showed no association with risk of hypertension [9], but a recent cohort found that raw vegetable intake was inversely associated with blood pressure (BP) and hypertension, while cooked vegetable intake was not [10]. A small randomized controlled trial (n = 87) revealed no effects on serum lipids during 8-week follow-up [11], similar to a large cross-sectional analysis that reported null association between vegetable intake and hyperglyceridemia [12]. Carter et al. reported an inverse association between vegetable intake with glycated hemoglobin (HbA1c) and OGTT [13], but a meta-analysis of cohort studies found no association with risk of type 2 diabetes [14].

So far, most of the evidences are observational or generated from small randomized trials with short follow-up periods. The observational associations are very likely to be limited by residual confounding, whereas the small trails with short follow-up periods provide restricted evidence for the long-term effects. Mendelian randomization is a study design that uses genetic variants -- usually single nucleotide polymorphism (SNPs) -- as instrumental variables to make causal inference for the exposure-outcome association, reflecting the overall effects of lifelong exposure [15]. It is analogous to randomized controlled trial in a way that genotypes are independently assorted during meiosis and fixed at fertilization, which enables MR to limit potential bias of residual confounding and reverse causality [15]. Furthermore, it remains unclear whether the potential association would differ between cooked and raw vegetable intake. Therefore, this study was aimed to evaluate the association between cooked and raw vegetable intake with metabolic risk factors using MR approach.

## 2. Methods

This two-sample MR study was reported based on STROBE-MR reporting guideline [16]. Summary-level data from genome-wide association studies (GWAS) were used for analysis, thus no specific ethic approval was required.

### 2.1 Genetic instrument selection

We identified genetic variants from three GWAS of cooked and raw vegetable intake [17–19]. In UK biobank, the intake of cooked and raw vegetable (number of heaped tablespoons; one heaped tablespoon is roughly equivalent to one serving) was measured at baseline via a touchscreen diet frequency questionnaire. The repeatability and validity of this diet questionnaire have been evaluated and confirmed in a previous study: the repeatability was 82% for cooked vegetable and 72% for raw vegetable in a repeat assessment after four years, and high agreement was observed when compared to 24-hour diet recall assessment [20]. Cooked and raw vegetable intake were weakly correlated in UK biobank, with a correlation coefficient of 0.30 [21].

We combined all SNPs that were associated with cooked or raw vegetable intake at genome-wide significance level (p < 5 * 10^−8^) in the three GWAS [17–19], and removed those that were duplicated, or rare variants (minor allele frequency < 1%) or in linkage disequilibrium. To further minimise potential horizontal pleiotropy, we searched Phenoscanner v2 database for any associated phenotypes for each SNP, and removed the SNPs that are associated with potential confounders, such as smoking, alcohol drinking, blood pressure and adiposity. The SNPs have been used in a previous study (unpublished). We searched their nearest genes and the biological functions for each SNP, and removed the SNPs that have regulatory effects on metabolism of lipids, glucose or protein. For the SNPs that could not be matched in outcome GWAS, we first tried to identify proper proxy SNPs in linkage disequilibrium (R^2^ > 0.8); if no proper proxy was identified, the unmatched SNPs were removed from further analysis (Supplementary figure 1).

We extracted the association estimates between the SNPs and vegetable intakes from the GWAS by Canela-Xandri et al [19], as this GWAS has a large sample size and adjusted for more covariates. It included 452264 unrelated European-ancestry individuals from UK Biobank, and adjusted for sex, age, age square, array batch, assessment centre and the 20 leading genetic principle components (Table 1). The strength of the genetic instruments was evaluated with F statistics, with F statistics > 10 suggesting good instrument strength [22]. The characteristics of the SNPs are shown in Table 2.

In total, we identified 9 and 19 eligible SNPs for cooked and raw vegetable intake, which explained 0.5% and 2.4% of phenotypic variance, respectively. The SNPs are located in different gene loci. The mechanisms behind the selected SNPs and cooked or raw vegetable consumption are suggested to be mediated by their regulations on olfactory receptors (for example, *rs9323534*), implying that smell and taste influence individual’s preference and choice on food [18].

### 2.2 Outcome data

The outcomes of interest included total cholesterol (TC), triglyceride (TG), LDL, high density lipoprotein cholesterol (HDL), BMI, SBP, diastolic blood pressure (DBP), pulse pressure (PP, the difference between SBP and DBP), fasting insulin (FI), fasting glucose (FG), glycated haemoglobin (HbA1c) and 2-hour glucose after oral glucose tolerance test (OGTT).

We used the summary-level GWAS statistics from Global Lipids Genetics Consortium (GLGC) for lipids-related outcomes (TC, TG, LDL, HDL) [23], Locke et al. 2015 for BMI [24], Meta-Analyses of Glucose and Insulin-related traits Consortium (MAGIC) for glycemic traits (FG, FI, OGTT, HbA1c) [25] and International Consortium of Blood Pressure (ICBP) for blood pressure (SBP, DBP, PP) [26], respectively. All these GWAS were conducted in unrelated European-ancestry individuals, and adjusted for sex, age, age square, genetic principle components and other study-specific covariates. (Table 1)

GLGC [23] included 188578 individuals who were not on lipid-lowering treatment, and the blood lipid levels were measured after > 8 hours of fasting. Locke et al.’s GWAS [24] included 339224 individuals. MAGIC [25] included 200622, 151013, 63396 and 146806 individuals for analyses of FG, FI, OGTT and HbA1c. FI was natural log-transformed. Participants were excluded if they had diagnosis of diabetes, or were on anti-diabetic medication, or had abnormal glycemic or insulin levels (FG > 7 mmol/L, OGTT > 11.1 mmol/L, HbA1c > 6.5%). ICBP [26] included 150134 individuals; for those on antihypertensive treatment, 15 mmHg was added to the raw SBP values (10 mmHg for DBP). BMI was additionally adjusted for in the GWAS of FG, FI, OGTT, SBP, DBP and PP. (Table 1)

### 2.3 Statistical analysis

Summary-level association statistics for each SNP were orientated across different GWAS so that their effect estimates were aligned on the same allele. We first performed univariable MR analysis with inverse variance weighted method, weighted median method and MR-Egger method. Inverse variance weight method assumes all genetic instruments are valid and no horizontal pleiotropy [27]. The median-based method assumes at least half of the genetic instruments are valid [28]. MR-Egger method can detect and correct for potential pleiotropy [29]. Pleiotropy was examined with MR-Egger intercept test, with a p value < 0.05 suggesting presence of directional pleiotropy, where MR-PRESSO method was used. MR-PRESSO method can detect outlier SNPs and provide effect estimates after removing the outliers [30]. Cochrane’s Q test was used to examine the heterogeneity across the genetic instruments, with p < 0.05 suggesting presence of heterogeneity and leave-one-out analysis would be performed to qualitatively evaluate the effect of heterogeneity on the association estimates.

Multivariable MR with inverse variance weighted method was performed as primary analysis to investigate the associations of cooked and raw vegetable intake with the outcomes of interest. Median-based method and MR-Egger method and were performed as sensitivity analyses.

Since all the outcomes were continuous variable, coefficient *beta* and their 95% confidence interval (CI) were used to quantify the association, indicating the change in the outcome for lifelong increase of one serving of vegetable intake. Bonferroni correction of significance level was used to control multiple comparison (0.05/(12*2) = 0.002). All analyses were performed using “*MendelianRandomization*” package (version 0.5.1), “*MR-PRESSO*” package (version 1.0) in R environment (version 4.1.1; R Core Team, Vienna Austria).

## 3. Results

The average F statistics was 30.53 (range 22.09 to 48.11) and 29.28 (range 10.95 to 48.82) for SNPs of cooked and raw vegetable intake, respectively, suggesting good instrument strength. (Table 2)

Similar null associations between vegetable intakes and the outcomes were observed in univariable MR, which was consistent across inverse variance weighted method, weighted median method and MR-Egger method (Supplementary table 1, Supplementary figure 2). Although MR-Egger intercept test suggested pleiotropy in the association between raw vegetable intake with BMI and FI, MR-PRESSO identified one (*rs6729029*) and zero outlier SNP, respectively, and removing the outlier did not change the results. Heterogeneity was observed in the association between cooked vegetable with TG, SBP and PP, and between raw vegetable with LDL, BMI and FG; however, in the leave-one-out analyses, the association estimates remained largely unchanged for most associations (Supplementary figure 3). For raw vegetable intake – FG association, *rs2447090* was likely to be an outlier, but removing it yielded association estimates of 0.05 (95% CI: 0.01, 0.10; p = 0.02), not passing the Bonferroni corrected significance level (Supplementary figure 3).

Overall in the primary multivariable MR with inverse variance weighted method, genetically determined vegetable intake was not associated with serum lipids, BMI, glycemic traits or BP (Figure 1, Supplementary table 2). For each serving increase of cooked vegetable intake, the *beta* for TC, BMI, FG and SBP were −0.06 (−0.30, 0.18; p = 0.62), −0.18 (−0.41, 0.05; p = 0.13), 0.01 (−0.11, 0.13; p = 0.84) and −2.43 (−6.36, 1.49; p = 0.23), respectively. For per serving increase of raw vegetable intake, the *beta* for TC, BMI, FG and SBP were −0.02(−0.20, 0.16; p = 0.84), 0.12 (−0.05, 0.29; p = 0.15), 0.03 (−0.06, 0.11; p = 0.56) and 0.94 (−1.91, 3.78; p = 0.52), respectively. A suggestive association between raw vegetable intake and reduced OGTT was observed (−0.33 [−0.64, −0.01; p = 0.04]), but it did not pass the Bonferroni corrected significance level. MR-Egger method and median-based method also showed similar results of null associations, and MR-Egger intercept test suggested no evidence of horizontal pleiotropy (Supplementary table 2). For FG, removing *rs2447090* in multivariable MR yielded −0.01 (−0.10, 0.10; p = 0.97) and 0.05 (−0.03, 0.12; p = 0.23) for cooked and raw vegetable intake, respectively, consistent with the primary analysis.

## 4. Discussion

In this MR study using summary level statistics from so far the largest eligible GWAS and the genetic variants as strong instrumental variables for cooked and raw vegetable consumption, we found no evidence for associations between vegetable intake with serum lipids, glycemic measures, BMI and BP, consistent across primary analysis and sensitivity analyses.

Previous evidence on the effects of vegetable intake on metabolic risk factors are mainly from observational studies or small randomized controlled trials, and showed substantial inconsistency. For serum lipids, a randomized crossover study of ten healthy individuals reported that a diet high in vegetable reduced serum LDL and HDL [31], similar to a randomized trial reporting that increasing vegetable intake from 94 g/day to 400 g/day for 3 weeks reduced TC, LDL, HDL and BMI among 60 overweight Asian women [32]. However, another randomized trial with a larger sample size (n = 76) showed no effects on serum lipids and lipoproteins during 8 weeks [11], consistent with a large cross-sectional study showing no association with hyperlipidemia [12]. For obesity, a meta-analysis of eight randomized trials of totally 1000 individuals showed that higher vegetable intake reduced body weight by 0.68 kg during a mean follow-up of 14 weeks [5], similar to a randomised trial of 250 individuals [33]; however, a recent meta-analysis of observational studies including 0.5 million individuals reported null associations with body weight outcomes [8]. For BP, a review of 8 observational studies found no association with risk of hypertension [9], echoed by a later study reporting null association with SBP or DBP neither cross-sectionally nor longitudinally [34]. For glycemic traits, a systematic review of 23 cohorts found no association with risk of type 2 diabetes [14]. A meta-analysis of nine studies found no association between vegetable intake and metabolic syndrome [35].

Causal inference has been difficult solely based on observational evidence and small trials, as they are prone to residual confounding, reverse causation and low statistical power. MR design is able to methodologically limit residual confounding and reverse causation [15]. Using the carefully selected SNPs as strong instruments for vegetable intake and the GWAS data of by far the largest sample size, this MR study is well powered to detect any clinically significant effect of vegetable intake.

There are three underlying assumptions for multivariable MR design. First, the genetic instrumental variables are associated with at least one of the exposures. Second, the genetic instruments are not associated with confounders of the exposure-outcome associations. Third, there are no independent pathways between the genetic instruments and the outcomes other than via one or more of the exposures [15]. For the first assumption, we selected the SNPs that are associated with cooked or raw vegetable intake at genome-wide association level of significance from three GWAS. Although the proportion of phenotype variance explained by the SNPs are relatively small (0.5% and 2.4%), which is not uncommon for behaviour phenotypes, the F statistics indicated their good instrumental strength. Biologically, the mechanism between genetics and vegetable intake is likely to be mediated by the regulatory effects of genes (for example, SNP *rs9323534* in gene *OR4K17*) on olfactory receptors [18]. These SNPs have been previously used [unpublished data]. For the second assumption, we removed the SNPs that are associated with smoking, drinking, obesity, BP, and metabolism of lipids, glucose and protein. The SNPs were not associated with sex, physical activity, red meat intake, and processed meat intake, as shown in a previous MR that constructed a polygenetic risk score using similar SNPs for vegetable intake [unpublished data]. For the third assumption, MR-Egger intercept test showed no evidence of horizontal pleiotropy in multivariable MR (Supplementary table 2). Therefore, the three assumptions are plausibly satisfied in our study.

Serum dietary-derived antioxidants, such as vitamin C and vitamin E, are valid biomarkers for vegetable intake [36]. Previously, these antioxidants have also been proposed to be the potential mediators for the protective effects of vegetable intake on metabolic risk factors [37], which, however, has been challenged by recent research. Although observational evidence suggested that serum vitamin C was associated with reduced BP [38,39], meta-analysis of trials found no effects of vitamin C supplementation on BP [40]. Furthermore, recent meta-analyses of randomized trials found that vitamin C supplementation was not associated with TC, TG, LDL, HDL [40–42], glucose, HbA1c and insulin [40,43], in line with the findings from multiple MR studies that circulating vitamin C level did not change lipids, BMI, BP [44] and glycemic traits [45]. For vitamin E, MR evidence showed that circulating vitamin E was not associated with BMI, or glucose traits [46]. Our findings of the null effects of vegetable intake are actually consistent with these recent trial and MR evidences on the antioxidants.

Since the metabolic risk factors are well established major risk factors for cardiovascular disease, the findings also imply limited evidence for a protective effect of vegetable intake on cardiovascular health. A previous study meta-analyzed one-sample and two-sample MR evidence and found null association between vegetable intake and coronary heart disease, stroke, heart failure and atrial fibrillation [unpublished]. Previous MR using serum antioxidants as exposures also showed similar results. For example, serum vitamin C was not associated with multiple cardiovascular diseases [44,47,48]; other antioxidants (vitamin E, carotene, lycopene and retinol) were not associated with risk of ischemic stroke [49] and CHD [50].

However, the findings should be interpreted carefully. The MR estimates suggest the change in outcomes for lifelong increase of one serving vegetable intake, assuming all other risk factors unchanged. Therefore, our findings should be interpreted as: solely increasing vegetable intake was very likely not to change the metabolic risk factors, while keeping fixed all other risk factors, including intakes of other food. In reality, diet is always complex with various kinds of food and substitutions between them. Given the relatively stable intake of total calorie, increasing vegetable intake is related to lower intake of other food [51,52]. The question is which food is replaced.

Therefore, in this sense, our finding is still in line with the recommendation for a balanced diet. There are some limitations with this study. First, we only included GWAS of European-ancestry individuals, so generalization to populations of other ethnicities may be limited. Second, the dietary pattern was measured in British population, thus reflecting a British diet and generalization to populations with a different dietary pattern should be cautious. Third, the SNPs explained only a small proportion of the phenotypic variances, which is common for behavioural trait, but F statistics suggested good instrument strength. Fourth, although we have differentiated cooked and raw vegetable intake, specific vegetable kinds and cooking methods (for cooked vegetable) may have specific effects, which warrants future research.

## 5. Conclusion

This two-sample MR study demonstrated null evidence for association between vegetable intake with serum lipids, BP, BMI and glycemic measures. The findings are consistent with previous genetic evidences of no association between vegetable intake with cardiovascular diseases, as well as of null association between diet-derived antioxidants with metabolic risk factors and cardiovascular diseases. This suggests that solely increasing vegetable intake has limited effects on metabolic risk factors.

## Data Availability

All data produced in the present study are available upon reasonable request to the authors

## Data availability

available upon request.

